# Taxonomic Signatures of Long-Term Mortality Risk in Human Gut Microbiota

**DOI:** 10.1101/2019.12.30.19015842

**Authors:** Aaro Salosensaari, Ville Laitinen, Aki Havulinna, Guillaume Meric, Susan Cheng, Markus Perola, Liisa Valsta, Georg Alfthan, Michael Inouye, Jeramie D. Watrous, Tao Long, Rodolfo Salido, Karenina Sanders, Caitriona Brennan, Gregory C. Humphrey, Jon G. Sanders, Mohit Jain, Pekka Jousilahti, Veikko Salomaa, Rob Knight, Leo Lahti, Teemu Niiranen

## Abstract

The collection of fecal material and developments in sequencing technologies have enabled cost-efficient, standardized, and non-invasive gut microbiome profiling. As a result, microbiome composition data from several large cohorts have been cross-sectionally linked to various lifestyle factors and diseases.^1–5^ In spite of these advances, prospective associations between microbiome composition and health have remained uncharacterized due to the lack of sufficiently large and representative population cohorts with comprehensive follow-up data.^6–8^ Here, we analyse the long-term association between gut microbiome variation and mortality in a large, well-phenotyped, and representative population cohort (*n* = 7211, FINRISK 2002; Finland).^9^ We report specific taxonomic and functional signatures related to the *Enterobacteriaceae* family in the human gut microbiome that predict mortality during a 15-year follow-up. These associations can be observed both in the Eastern and Western Finns who have differing genetic backgrounds, lifestyles, and mortality rates.^10,11^ Our results supplement previously reported cross-sectional associations,^1–4,12^ and help to establish a methodological and conceptual basis for examining long-term associations between human gut microbiome composition, incident outcomes, and general health status. These findings could serve as a solid framework for microbiome profiling in clinical risk prediction, paving the way towards clinical applications of human microbiome sequencing aimed at prediction, prevention, and treatment of disease.

---

The long research tradition in population-level health surveys, high participation rates, and the availability of comprehensive, nationwide health registers that allow monitoring of health variations across an individual’s life span have brought Finland to the forefront of population-based cohort studies.^9,13–15^ Here, we analyze the fecal microbiome composition in a representative random sample of 7211 adults (mean age 49.5 years, 55.1% women) who participated in the FINRISK 2002 population survey which included stool sample collection and cross-sectional phenotyping in 2002 (**Fig. 1a–b**).^9^ Access to electronic health registers and death certificates across the 15-year time span following sample collection is a unique feature of this study allowing us to complement the earlier cross-sectional studies by associating gut microbiome profiles with a long-term follow-up of health status and mortality after the baseline examination. Altogether, 729 of the 7055 participants (10.2%) with complete data available died during a median follow-up of 14.2 years.

**Figure 1.**
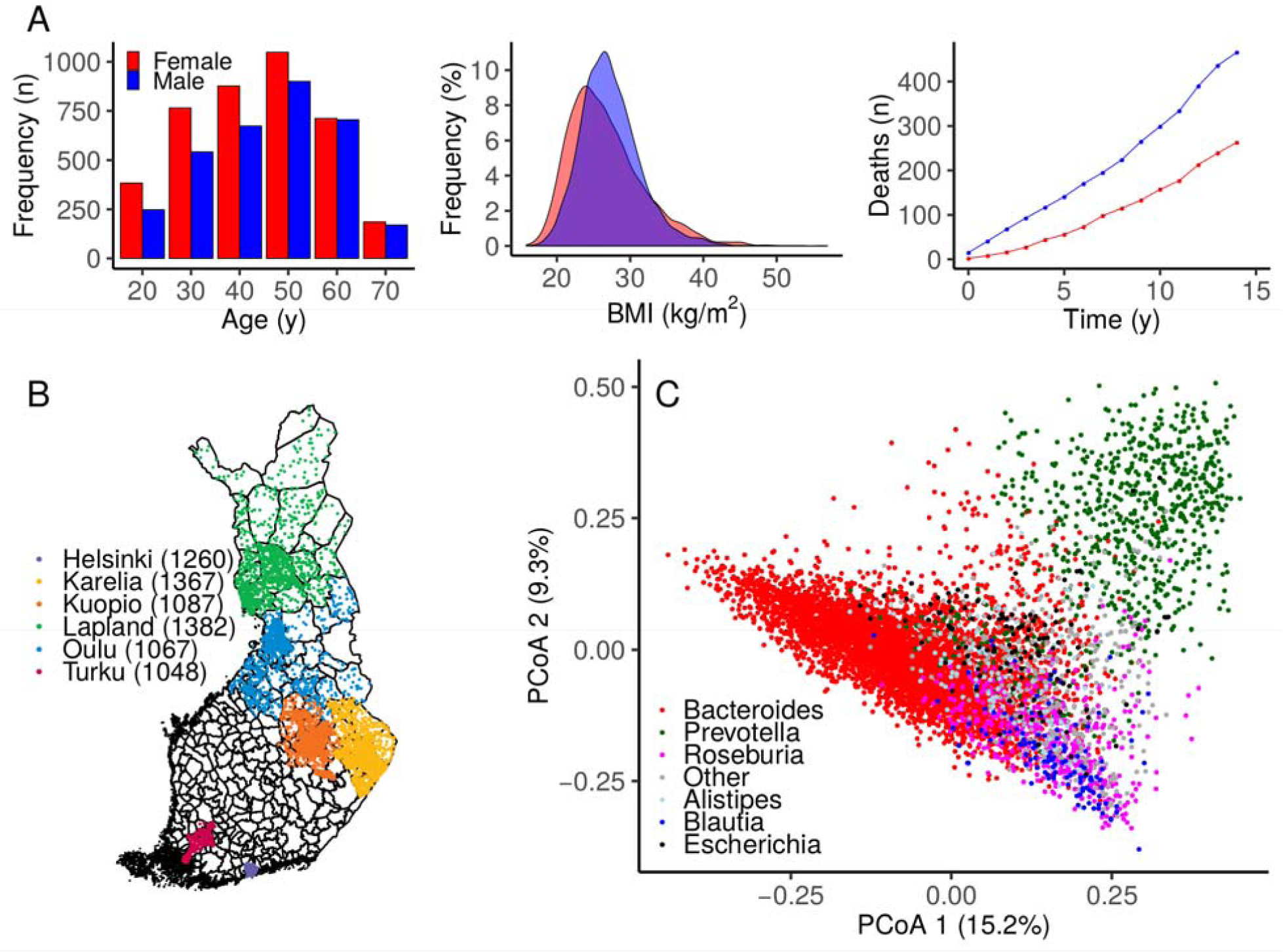
Study sample and gut microbiome characteristics. **A** At baseline, the study sample (N=7211) had a balanced sex ratio (55% women), a mean age of 49 years (range 24-74; left panel), and a mean body mass index (BMI) of 27 kg/m^2^ (range 16-57; middle panel). During a median follow-up time of 14.2 years, 721 of 7055 (10.2%) participants with complete data who were included in the prospective analysis died (right panel). **B** A total of 7211 out of 13500 randomly sampled individuals (53.4% participation rate) from six catchment areas in Finland underwent stool sampling, a physical examination, and filled in a questionnaire on health behaviour, history of diseases and current health. **C** Principal coordinates analysis (PCoA) indicates sample similarity based on species-level taxonomic composition. The colour indicates the dominant genus in each sample. Altogether 96% of the samples are dominated by one of the most common dominant genera that are indicated in the figure.

We investigated links between mortality and the key features of microbiome composition, including alpha and beta diversity, core microbiota, and taxonomic co-occurrence networks. We identified a total richness of 67 phyla and 2019 genera (**Supplementary Tables 1–2**). Interindividual variation in taxonomic composition was largely attributable to differences in the relative abundances of the most prevalent and abundant core groups (**Fig. 1c; Extended Data Fig. 1; Supplementary Table 2**). Most of the groups were rare and observed in <1% of the study population. In addition to overall species diversity, we have focused on the core microbiome that comprised the 95 genus-level taxonomic groups that exceeded the within-sample relative abundance of >0.1% in >1% of samples. These included mostly bacterial genera (87) and plasmids (4) but also viruses (1) and archaea (3) (**Supplementary Table 2**). The median relative abundance of the combined core groups was 99.3%.

We performed a prospective analysis by examining how microbiome features predicted mortality risk in a 15-year follow-up. Alpha diversity was not a significant predictor of mortality (**Supplementary Table 3**). However, we detected a robust and significant signal between beta diversity, or the overall community variation as measured by Aitchison distance, and elevated mortality risk. Namely, the third principal component of CLR-transformed species abundance matrix (PC3) was a strong predictor of all-cause mortality (**Fig. 2a–b; Supplementary Table 3**). The observation was robust to factors known to affect microbiome composition and mortality risk, i.e., age, sex, BMI, smoking, diabetes, use of antineoplastic or immunomodulating agents, systolic blood pressure, and use of antihypertensive medication. Moreover, the findings related to PC3 could be observed in independent samples of the Eastern and Western Finnish populations whose genetic backgrounds, lifestyles, and life expectancies differ (**Extended Data Fig. 2**).^10,11^ PC3 was driven by species of the *Enterobacteriaceae* family that is a part of the normal gut microbiome but is known to cause infectious diseases in other body sites **Extended Data Figs. 3 and 4; Supplementary Table 4**).^16^ Several *Enterobacteriaceae* genera also individually predicted mortality (**Supplementary Table 5**). There was limited statistical power for detecting association between mortality and enrichment of virulence genes as their prevalence among the FINRISK samples was low (<1%; **Supplementary Table 6**). These genes were mainly related to virulent strains of *E. coli*.

**Figure 2.**
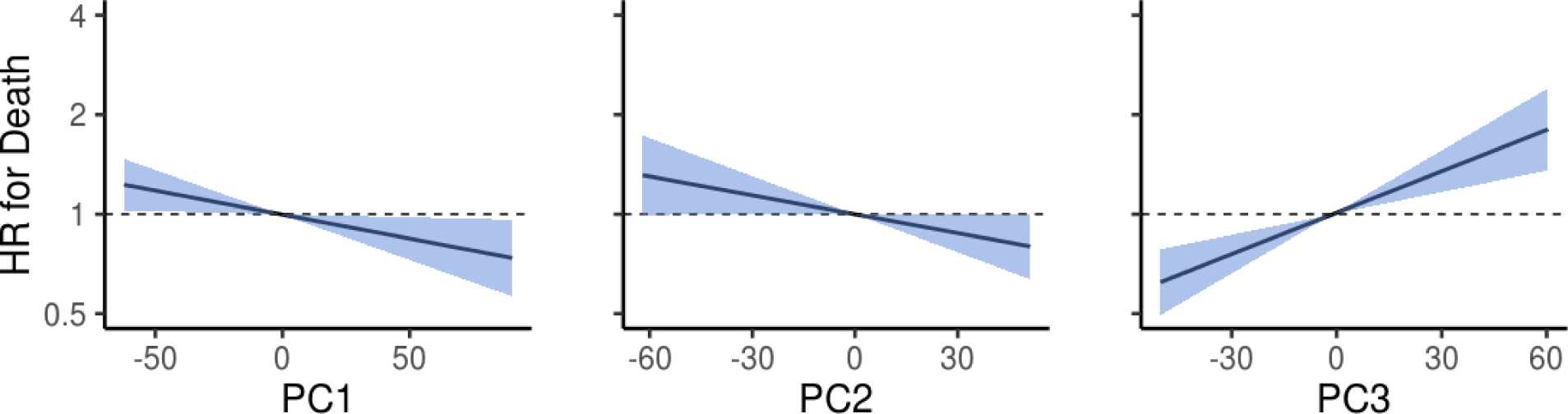
Microbiome principal components as predictors of death. Association between the first three principal components (PC) and mortality risk. Blue area indicates the 95% confidence interval (CI) of the hazard ratio. Unit variance increase in the PCs were related to hazard ratios of 0.918 (95% confidence interval [CI], 0.852–0.989; FDR-adjusted *P* = 0.0505), 0.933 (95% confidence interval [CI], 0.869–1.001; FDR-adjusted *P* = 0.072), 1.155 (95% confidence interval [CI], 1.077–1.239; FDR-adjusted *P* = 2.1×10^−4^) for PC1 - PC3, respectively. Analyses are adjusted for age, body mass index, sex, smoking, diabetes, use of antineoplastic and immunomodulating agents, systolic blood pressure and self-reported antihypertensive medication.

We then investigated the overall capacity of taxonomic composition in predicting mortality risk. After identifying significant linear and nonlinear associations between the abundances of 43 genera and mortality (**Extended Data Fig. 5a; Supplementary Table 5**), we applied a random survival forest model to identify a taxonomic signature that is maximally predictive of all-cause mortality (**Extended Data Fig. 3b; Supplementary Table 7**). The top predictors in this supervised analysis also included multiple *Enterobacteriaceae* genera (**Extended Data Fig. 5b**). However, community composition did not improve mortality prediction compared to the covariates (C-statistic 0.798 for covariates versus 0.794 for covariates plus community composition; *P* = 0.049. C-statistics for the community composition alone was 0.633).

In order to pinpoint specific taxonomic markers that could predict mortality risk, we complemented the community-level analyses by shifting the focus towards more refined sub-community analysis. We identified groups of tightly clustered genera based on taxonomic co-occurrence network analysis. All four strongest network modules, or subnetworks (**Fig. 3; Extended Data Fig. 6**), included genera that were predictors of all-cause mortality. We observed the strongest intra-network correlations and mortality associations for the subnetwork that consisted mainly of *Enterobacteriaceae* genera (**Fig. 3b**). This subnetwork was observed in both Western and Eastern Finnish populations (**Extended Data Fig. 7)**. The total abundance of this subnetwork predicted mortality (hazard ratio = 1.15, 95% confidence interval [CI], 1.07– 1.23; *P* = 0.010).

**Figure 3.**
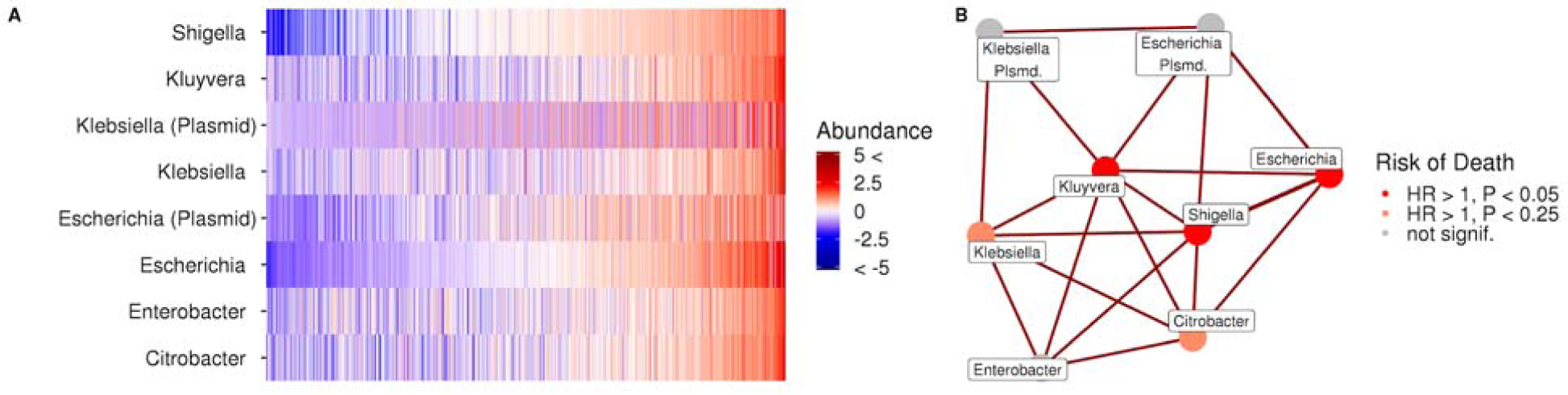
Microbial subnetworks, community structure, and their relation to mortality. **A** Abundance variation across the study population for the subnetwork that exhibits the strongest mortality associations (CLR-transformed abundances centered at zero and scaled to unit variance). The samples are ordered by the total relative abundance of the subnetwork. **B** The observed subnetwork structure and mortality risk. The total subnetwork abundance was associated with mortality with a hazard ratio of 1.15 (95% confidence interval [CI], 1.07–1.24; *P* = 0.012). The respective hazard ratios were 1.17 (95%, 1.08–1.27; *P* = 0.027) in the Eastern and 1.11 (95%,0.98–1.27; *P* = 0.607) in the Western Finnish populations. The analyses are conducted for microbial core and adjusted for age, body mass index, sex, smoking, diabetes, use of antineoplastic and immunomodulating agents, systolic blood pressure and self-reported antihypertensive medication; *P* values are FDR-adjusted.

Using Kyoto Encyclopedia of Genes and Genomes (KEGG) orthology groups, we assessed the potential microbial functional roles in individuals with an elevated mortality risk (**Fig. 4**; **Extended Data Fig. 8; Supplementary Table 8**). Many mortality-associated markers were involved in the KEGG categories related to drug biodegradation, carbohydrate metabolism, lipid metabolism, and infectious diseases. These associations were both positive and negative (**Fig. 4**; **Extended Data Fig. 8; Supplementary Table 8**). Numerous prior studies have demonstrated that gut microbes can affect lipid and glucose metabolism and their circulating levels which, in turn, may affect the risk of cardiometabolic disease.^17,18^ Furthermore, it has been previously demonstrated that the gut microbiota can exert direct effects on drug metabolism, potentially affecting disease risk through drug efficacy and toxicity.^19^ Functional pathways that were negatively associated with mortality also included biological processes related to the nervous system (**Extended Data Fig. 8)**. These functional predictions support our findings at the taxonomic level, while also suggesting that gut-drug-interactions, gut microbiome-metabolome interactions, and the gut-brain axis could play a role in the development of disease.^20,21^

**Figure. 4.**
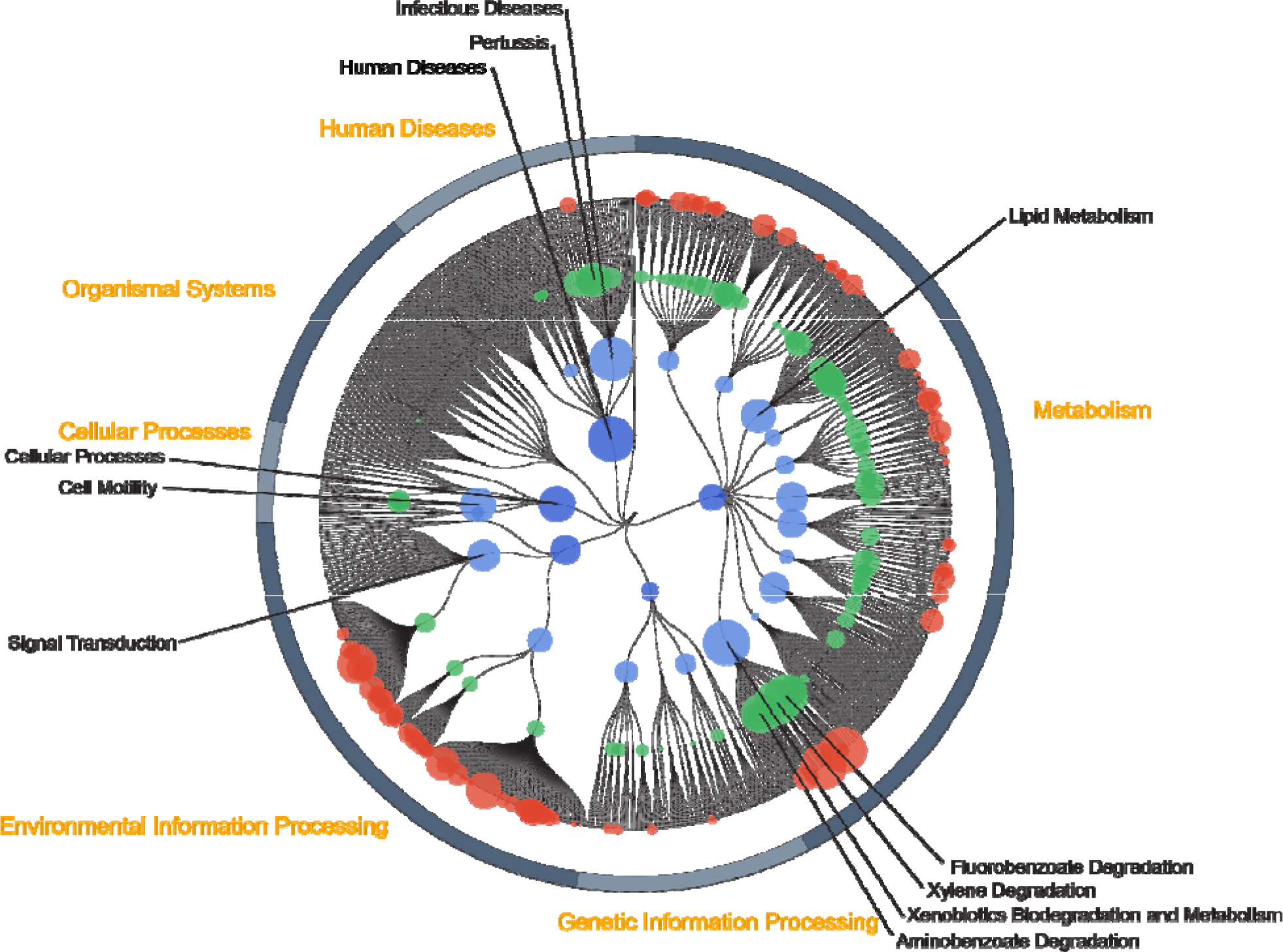
Functional pathways associated with increased risk of death. For the module (red), pathway (green), biological process (light blue), and biological category (dark blue) functional layers, node size corresponds to the average inverse *P* value of the KEGG Orthology group assigned to that node. Only KO groups that were positively associated with mortality were included. Node titles are shown for nodes in the three highest layers with a size > 200.

Our analysis provides a systematic quantification of the long-term health associations of the human fecal microbiome. In spite of using a remarkably heterogeneous outcome variable (all-cause mortality), we could identify specific gut microbiome features that predicted all-cause mortality during the 15-year follow-up. However, despite being a heterogeneous outcome, all-cause mortality is also a robust end point as it is virtually free of misclassification or loss to follow-up. All deaths in Finland are registered in a nationwide database, except for a fraction of the rare cases where a person permanently moves abroad. The observed associations suggest that specific taxonomic configurations of the human gut microbiome may reflect health-associated changes that are linked to increased mortality, or potentially play a unique role in the maintenance of health and development of incident disease.^5,16,20^

In particular, we observed a robust link between mortality and several species and genera of the *Enterobacteriaceae* family. In prior cross-sectional human studies, *Enterobacteriaceae* have been observed to be enriched in patients with inflammatory bowel disease and colorectal cancer.^22^ It has been speculated that *Enterobacteriaceae*, normally dominant in the upper gastrointestinal tract, become enriched in the stool due to a faster stool transit time that occurs in diarrhea, a symptom of many gastrointestinal diseases.^23^ Host-mediated inflammation has also been shown to disrupt the gut microbiome and promote the overgrowth of *Enterobacteriaceae*.^24^ On the other hand, increased prevalence and enhanced virulence potential of gut *E. coli* have both been linked to urinary tract infections.^25^ In fact, extraintestinal *E. coli* strains often exist in the gut without consequences but have the capacity to disseminate and colonize other host niches including the blood, the central nervous system, and the urinary tract, resulting in disease.^26,27^

A particular strength of our analysis is the availability of a random population sample comprising thousands of adults from a homogeneous northern European population and the access to comprehensive electronic health registers. As such, this can complement the findings from earlier cross-sectional population studies that have had a more limited representativeness based on their focus on specific populations,^1,28^ lack of random sampling,^1–4^ or low participation rates.^29^ Although our data lacked certain covariates that have been recently linked to microbiome composition, such as stool consistency and fecal chromogranin A,^1–4^ our findings are based on a representative population sample, and the findings were supported in both Eastern and Western Finnish populations. Despite the lack of cohorts that could be used for external replication of our prospective results, our findings have implications on the design of future studies that aim to map microbiome-health associations across extended periods of time.

Until now, prospective long-term data linking microbiome composition with incident outcomes have been unavailable. Our data provide a proof-of-concept that the microbiome can be used to assess the overall mortality risk, and potentially for disease risk assessment. Additional studies will be needed to assess which disease states can be most effectively predicted through microbiome profiling. In addition, our findings can help establish a framework for recruiting disease-susceptible individuals to randomised trials to assess causal effects of gut microbiota variation on health outcomes. However, extensive research is still warranted before human microbiome sequencing can be used for prediction, prevention, and targeted treatment of disease.

## Methods

### Study sample

The FINRISK population surveys have been performed every 5 years since 1972 mainly to monitor trends in cardiovascular disease risk factors in the Finnish population. The FINRISK 2002 study was based on a stratified random sample of the population aged 25–74 years from specific geographical areas of Finland (**Fig. 1)**.^9^ The survey included participants from North Karelia and Northern Savo in eastern Finland, Turku and Loimaa regions in southwestern Finland, the cities of Helsinki and Vantaa in the capital region, the provinces of Northern Ostrobothnia in northwestern Finland, Kainuu in northwestern Finland and the province of Lapland in northern Finland. The sampling was stratified by sex, region and 10-year age group so that each stratum had 250 participants. In North Karelia, Lapland and the cities of Helsinki and Vantaa, the strata with 65–74 year old men and women were also sampled, each with 250 participants. The original population sample was thus 13500 (minus 64 who had died or moved away between sampling and the survey); the overall participation rate was 65.5% (*n* = 8798). The sampling has been previously described in detail.^9^ We successfully performed stool shotgun sequencing in *n* = 7231 individuals. After excluding 20 individuals with low read counts (<50000), *n* = 7211 participants (mean age 49.5 years, 55.1% women) remained for unsupervised analysis, of whom *n* = 7055 had the full covariate information available for survival analysis (**Fig. 1**). The study protocol of FINRISK 2002 was approved by the Coordinating Ethical Committee of the Helsinki and Uusimaa Hospital District (Ref. 558/E3/2001). All participants signed an informed consent. The study was conducted according to the World Medical Association’s Declaration of Helsinki on ethical principles. Due to a lack of external cohorts with microbiome and long-term mortality data, we used two internal subsamples of 4979 Eastern Finns (mainly from Northern Karelia, Northern Savo, Kainuu, and Northern Ostrobothnia regions) and 2232 Western Finns (mainly from Helsinki, Turku, and Loimaa regions). Altogether 4871 and 2184 samples had complete covariate information, respectively. We used this to examine the robustness of the results in distinct subpopulations within the cohort. These two subsamples were chosen due to their well-known differences in genetic backgrounds, lifestyles, and mortality rates.^10–12^

### Baseline examination

The FINRISK 2002 survey included a self-administered questionnaire, physical measurements and collection of blood and stool samples. The questionnaire, together with an invitation to the health examination, was sent by mail to all subjects. Trained nurses carried out a physical examination and blood sampling in local health centres or other survey sites. The participants were advised to fast for ≥4 hrs and avoid heavy meals earlier during the day. The venous blood samples were centrifuged at the field survey sites, stored in −70°C, and transferred daily to the laboratory of the Finnish Institute for Health and Welfare. Data for physiological measures, biomarkers, dietary factors, demographic factors, and lifestyle factors was collected. Details on the methods have been previously described.^9^

### Stool Sample Collection

At the baseline examination, all willing participants were given a stool sampling kit with detailed instructions. The participants mailed their samples overnight between Monday and Thursday under Finnish winter conditions to the laboratory of the Finnish Institute for Health and Welfare where the samples were stored at −20°C. The stool samples were stored unthawed until they were transferred in 2017 to the University of California San Diego for microbiome sequencing.

### Stool DNA extraction and library preparation

A miniaturized version of the Kapa HyperPlus Illumina-compatible library prep kit (Kapa Biosystems) was used for library generation, following the previously published protocol.^30^ DNA extracts were normalized to 5 ng total input per sample in an Echo 550 acoustic liquid handling robot (Labcyte Inc). A Mosquito HV liquid-handling robot (TTP Labtech Inc was used for 1/10 scale enzymatic fragmentation, end-repair, and adapter-ligation reactions). Sequencing adapters were based on the iTru protocol,^31^ in which short universal adapter stubs are ligated first and then sample-specific barcoded sequences added in a subsequent PCR step. Amplified and barcoded libraries were then quantified by the PicoGreen assay and pooled in approximately equimolar ratios before being sequenced on an Illumina HiSeq 4000 instrument to an average read count of approximately 900,000 reads per sample.

### Taxonomic and functional profiling from sequencing data

We analysed shotgun metagenomic sequences using a pipeline built with the Snakemake bioinformatics workflow library.^32,33^ We trimmed the sequences for quality and adapter sequences using Atropos,^34^ and removed host reads by genome mapping against the human genome assembly GRCh38 with Bowtie2.^35^ We assigned sequences taxonomy using SHOGUN v1.0.5^36^ against a database containing all complete bacterial, archeal, and viral genomes available from NCBI RefSeq as of version 82 (May 8, 2017). We processed the results to estimate the relative abundance of taxa. Functional profiles were calculated from a combination of observed and predicted KEGG Orthology group (KO) annotations from the RefSeq genomes following the default parameters of the SHOGUN tool^36^. Briefly, the final KO table represents a weighted average of directly observed functional genes and those estimated to be present but unsampled based on their predicted presence within an observed genome. A full description of the method has been published.^36^

### Virulence genes

The 7211 FINRISK samples were matched to the Virulence Factor Database (VFDB; DNA sequences of the full database).^38^ Anvi’o (v5.5) was used to build a Bowtie2 database from the VFDB FASTA files and to map the FINRISK reads to the VFDB genes using the Anvi’o default setting and 99% sequence similarity.^39^ The coverage was analyzed with samtools (v1.9). A coverage of 500bp and 90% of the VFDB gene length was required. The prevalence of the VFDB genes accepted with these filters is shown in **Supplementary Table 6**.^40^

Register linkage for pre-existing (prevalent) diseases, medication use at baseline, and mortality In Finland, each permanent resident is assigned a unique personal identity number at birth or after immigration, which ensures reliable linkage to the computerised health registers. The nationwide Finnish health registers ensure in practice 100% coverage of all major health events (Hospital Discharge Register), all prescription drug purchases (Drug Purchase and Special Drug Reimbursement Registers), and all deaths (Causes-of-Death Register). The quality of the diagnoses in the Finnish national registers has been previously validated.^13,14^ We obtained dates and causes of deaths from the national Causes-of-Death Register. The participants were followed through Dec. 31, 2016. We observed 729 deaths between the baseline and end of follow-up period; 519 deaths occurred in Eastern Finns and 210 in Western Finns (511 and 210 with complete covariate information).

### Covariates

We calculated body mass index as weight in kilograms divided by height in metres squared. Smoking (N=1648) was defined as daily use of tobacco products. We defined diabetes (N=401) as having a health event with an International Classification of Diseases code^41^ of E10-14 in the Hospital Discharge Register or having a drug reimbursement code for diabetes in the Special Drug Reimbursement Register prior to baseline. We defined systolic blood pressure based on the mean of three measurements performed by a nurse using a mercury sphygmomanometer. The participants self-reported antihypertensive medication use (N=1096). Use of antineoplastic or immunomodulating agents (N=62) was defined as a purchase of medications with an Anatomical Therapeutic Chemical code of L recorded in the Drug Purchase Register up to four months prior to baseline.^42^

### Statistical Methods

We conducted all statistical analyses using R^43^. In all association analyses, we standardised all phenotype variables except dichotomous variables. We controlled all cross-sectional and prospective association analyses for age, body mass index, sex, smoking, diabetes, use of antineoplastic or immunomodulating agents, systolic blood pressure and self-reported antihypertensive medication, unless otherwise indicated. We corrected for multiple testing using False Discovery Rate correction (Benjamini-Hochberg).^44^ We report the P-values, where we considered an FDR-corrected *P* < 0.05 significant.

### Core microbiota

We determined the 1% microbial core at the 0.1% detection limit (i.e., the taxa prevalent at least in 1% of the samples at >0.1% relative abundance).

### Principal co-ordinates and component analyses of microbiome variation

We calculated a Principal Coordinates Analysis (PCoA) of Bray-Curtis beta diversity matrix using compositional microbial species-level abundance data with the R package *phyloseq*^*45*^). We also analysed the beta diversity characterized by between-samples Aitchison distance by performing a Principal Component Analysis (PCA) using CLR-transformed species-level abundances (R function “prcomp”).

### Alpha diversity

We characterised the alpha diversity of the microbiome with the Shannon index using species-level abundance data.

### Subnetwork detection

We detected sparse taxonomic co-occurrence subnetworks with SPIEC-EASI^46^ (R package *SpiecEasi*) with default parameters and the (bounded) StARS model selection (“bstars”). We carried out StARS with the prior regularisation beta set at the recommended threshold.^47^ We excluded subnetworks with fewer than three members.

### Survival Analysis

We tested the association between CLR abundance of each genus and all-cause mortality using Cox proportional hazards models^48^ (two-tailed Wald tests for the coefficient corresponding to the genus; R package *survival*^*49*^). Moreover, to recover potential non-linear associations, we modelled genus abundance both linearly and with penalised cubic splines (R function “pspline” with the default parameters). The alpha diversity and principal components 1–5 were treated similarly. We assessed the proportional hazards assumption using Schoenfeld residuals.

We tested the relation of the core genera with mortality using multivariate random survival forest^50^ (R package *randomForestSRC*^*51*^). We used default settings and measured the predictor sets’ performance with Harrell’s C-statistic^52^ in 5-fold cross-validation and then calculated the importance scores using all subjects.

### Functional Analysis

We associated each KO group with mortality in Cox proportional hazards models. The KO groups were log(1+x)-transformed and analyzed using a standard linear model to determine the direction of association for all KO groups. We used the FuncTree application to analyse and visualise the functional enrichment of the gut microbiome in individuals with at an increased risk of death.^53^ For the module, pathway, and biological process layers, we used node sizes that corresponded to the average inverse *P* value of all KO groups that could be assigned to that node. Analyses were performed separately for KO groups that were related positively or negatively with mortality.

### Data availability

The data that support the findings of this study are available from the FINRISK Data Access Committee at Finnish Institute for Health and Welfare based on reasonable request [contact details available from T.N.]. The data are not publicly available due to them containing information that could compromise research participant privacy/consent.

### Code availability

The source code for the analyses will be shared with a permanent DOI from Zenodo.

## Data Availability

The data that support the findings of this study are available from the FINRISK Data Access Committee based on reasonable request [contact details available from T.N.]. The data are not publicly available due to them containing information that could compromise research participant privacy/consent.

## Supplementary information

Supplementary information is available in the online version of the paper.

## Acknowledgements

We thank all participants of the FINRISK 2002 survey for their contributions to this work, and Tara Schwartz for assistance with laboratory work. This research was supported in part by grants from the Finnish Foundation for Cardiovascular Research, the Emil Aaltonen Foundation, the Paavo Nurmi Foundation, the Urmas Pekkala Foundation, the Finnish Medical Foundation, the Academy of Finland (295741, 307127 to L.L.; 321351 to T.N.), UTUGS graduate school (to V.L.), and the National Institutes of Health (R01ES027595 to M.J., K01DK116917 to J.D.W., R01HL134168, R01HL131532, R01HL143227, and R01HL142983 to S.C.). Additional support was provided by Illumina, Inc. and Janssen Pharmaceutica through their sponsorship of the Center for Microbiome Innovation at UCSD.

## Author contributions

V.S., R.K, L.L and T.N, designed the work, A.H., J.W., M.P., L.V., G.A., G.M., T.L., M.J., P.J., V.S., R.S., K.S., C.B., and G.H. acquired the data, A.S., V.L., L.L., J.S., and T.N. analysed the data; and M.I., M.J., P.J., S.C., V.S., R.K., L.L, and T.N supervised the work. All authors wrote the manuscript and gave final approval of the version to be published.

## Author information

Reprints and permissions information is available at www.nature.com/reprints. Readers are welcome to comment on the online version of the paper. Correspondence and requests for materials should be addressed to T.N. and L.L.

## Extended data

**Extended Data Fig. 1.**
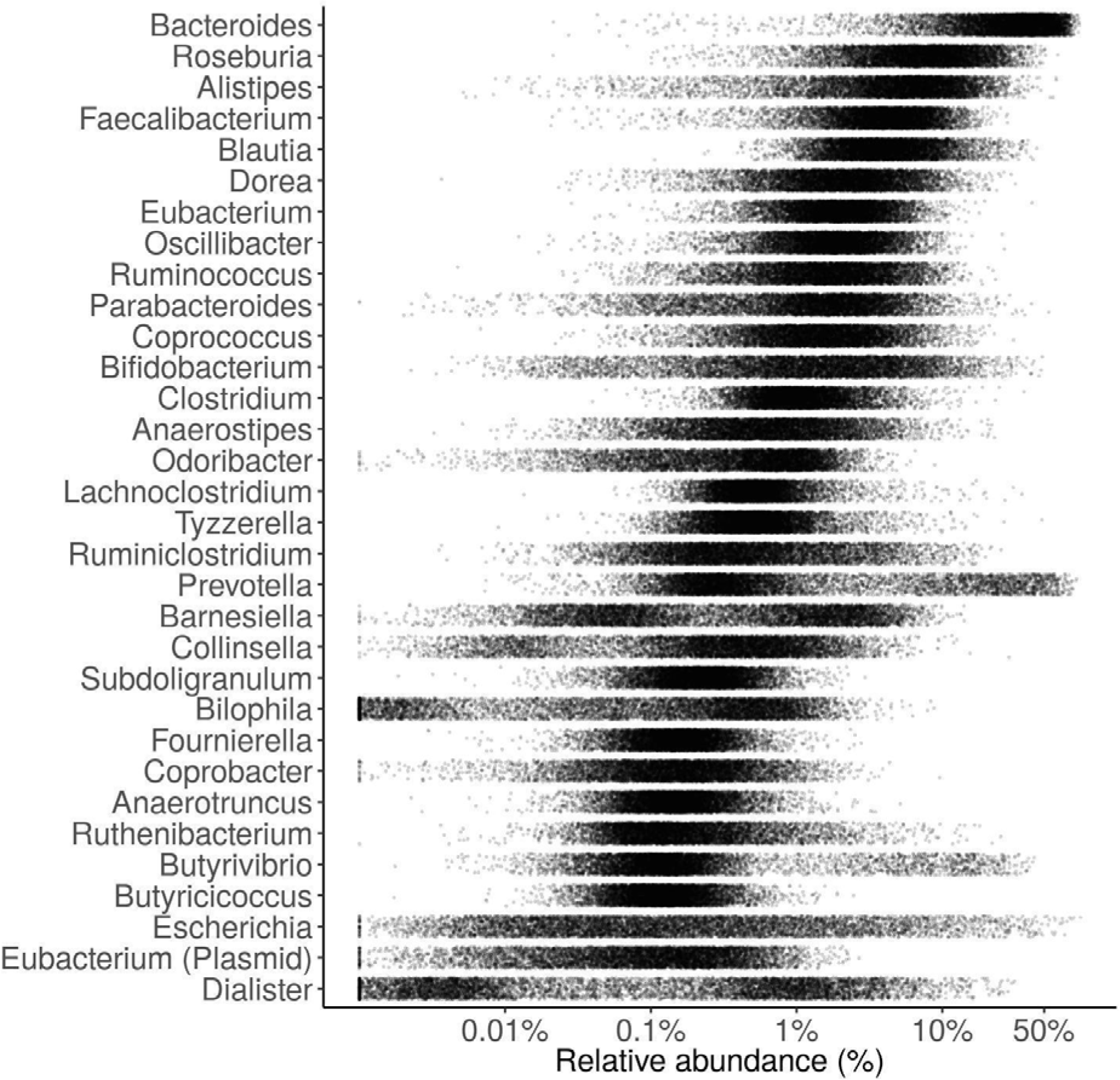
Relative abundances for the 32 most abundant and prevalent core genera that were detected at >0.1% relative abundance in the majority (>50%) of individuals in the study cohort. Each dot represents one individual, and the darker regions indicate more populated areas of the abundance landscape. On average, these genera cover 93.2% of the community based on their combined relative abundance.

**Extended Data Fig. 2.**
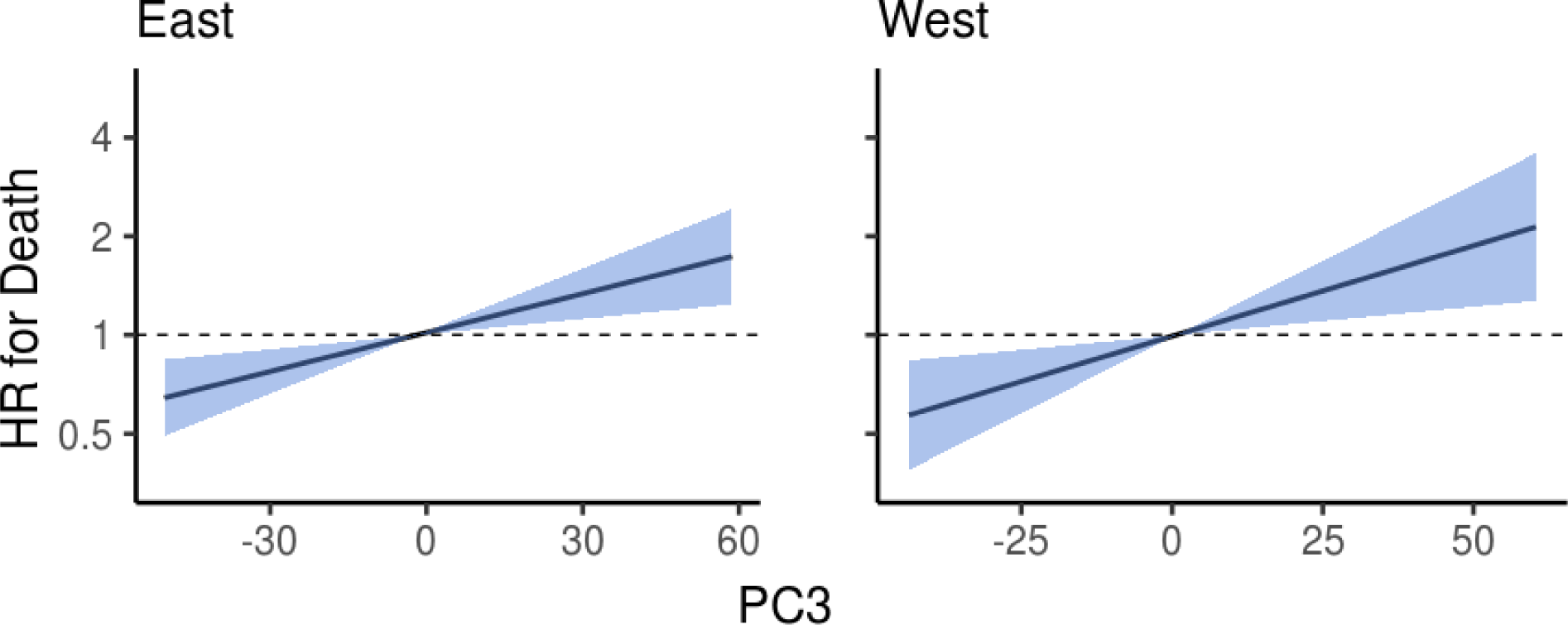
Association between the third principal component (PC3) and mortality in the Eastern and Western Finnish populations. Blue area indicates the 95% confidence interval (CI) of the hazard ratio. Unit variance increase in PC3 was related to hazard ratios of 1.147 (95% confidence interval [CI], 1.055–1.246; FDR-adjusted *P* = 2.6×10^−3^) and 1.207 (95% CI, 1.061–1.372; FDR-adjusted *P* = 4.1×10^−3^) in Eastern and Western Finnish populations, respectively. Analyses are adjusted for age, body mass index, sex, smoking, diabetes, use of antineoplastic and immunomodulating agents, systolic blood pressure and self-reported antihypertensive medication.

**Extended Data Fig. 3.**
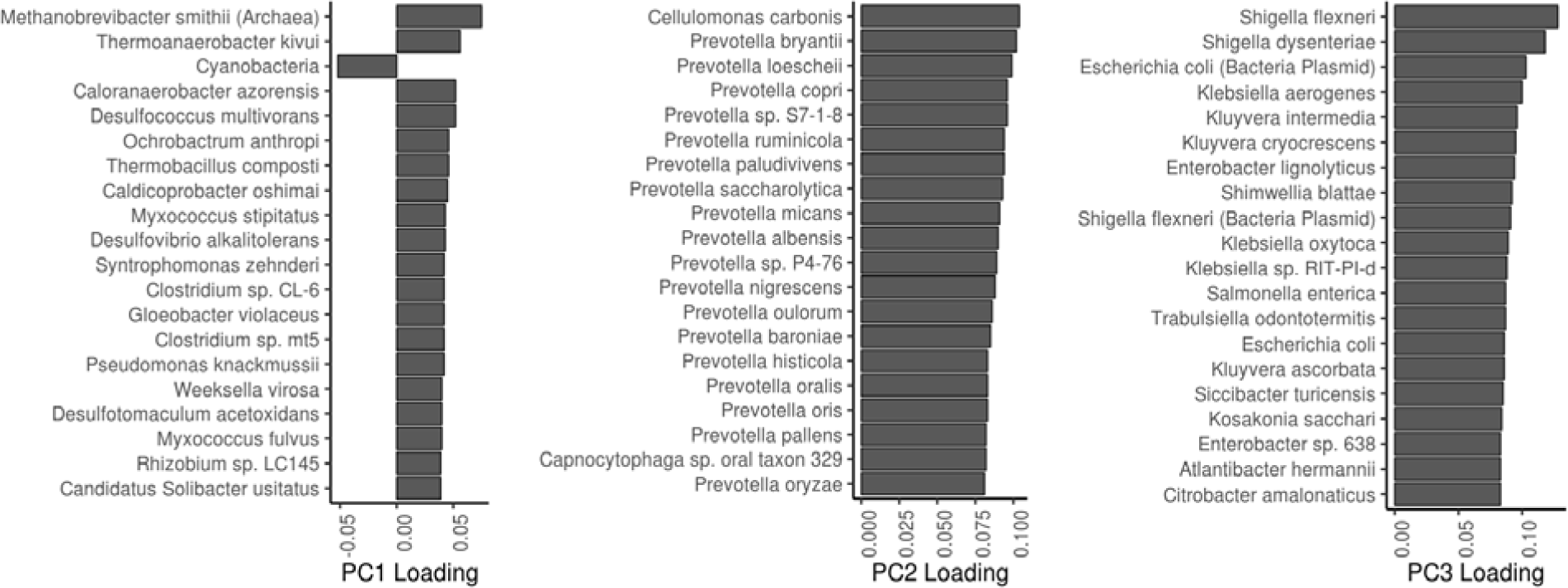
Principal component driver species. The 20 most important driver species of the first three principal components.

**Extended Data Fig. 4.**
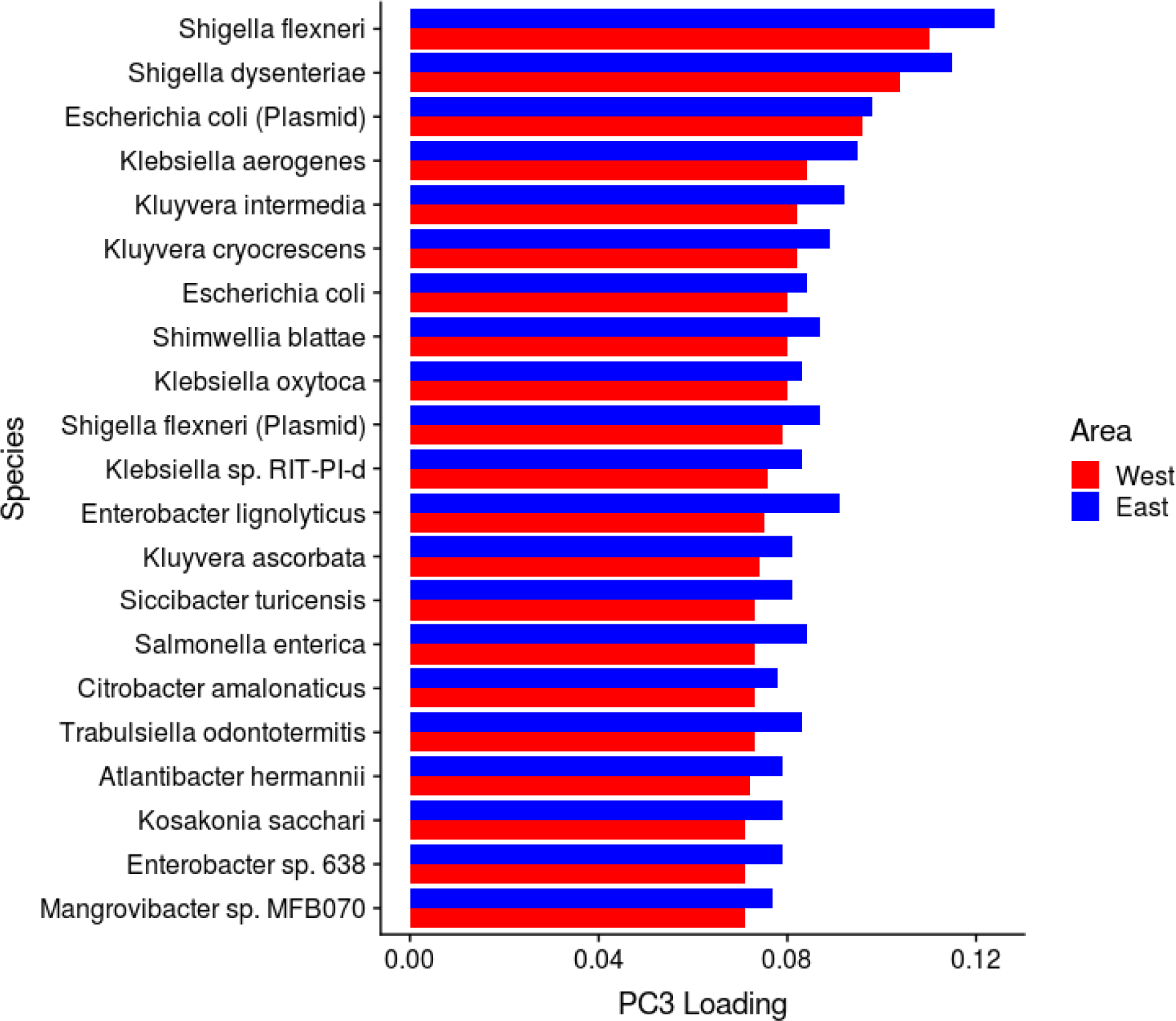
Third principal component drivers in Eastern and Western Finland. Most important PC3 drivers in Eastern and Western Finland.

**Extended Data Fig. 5.**
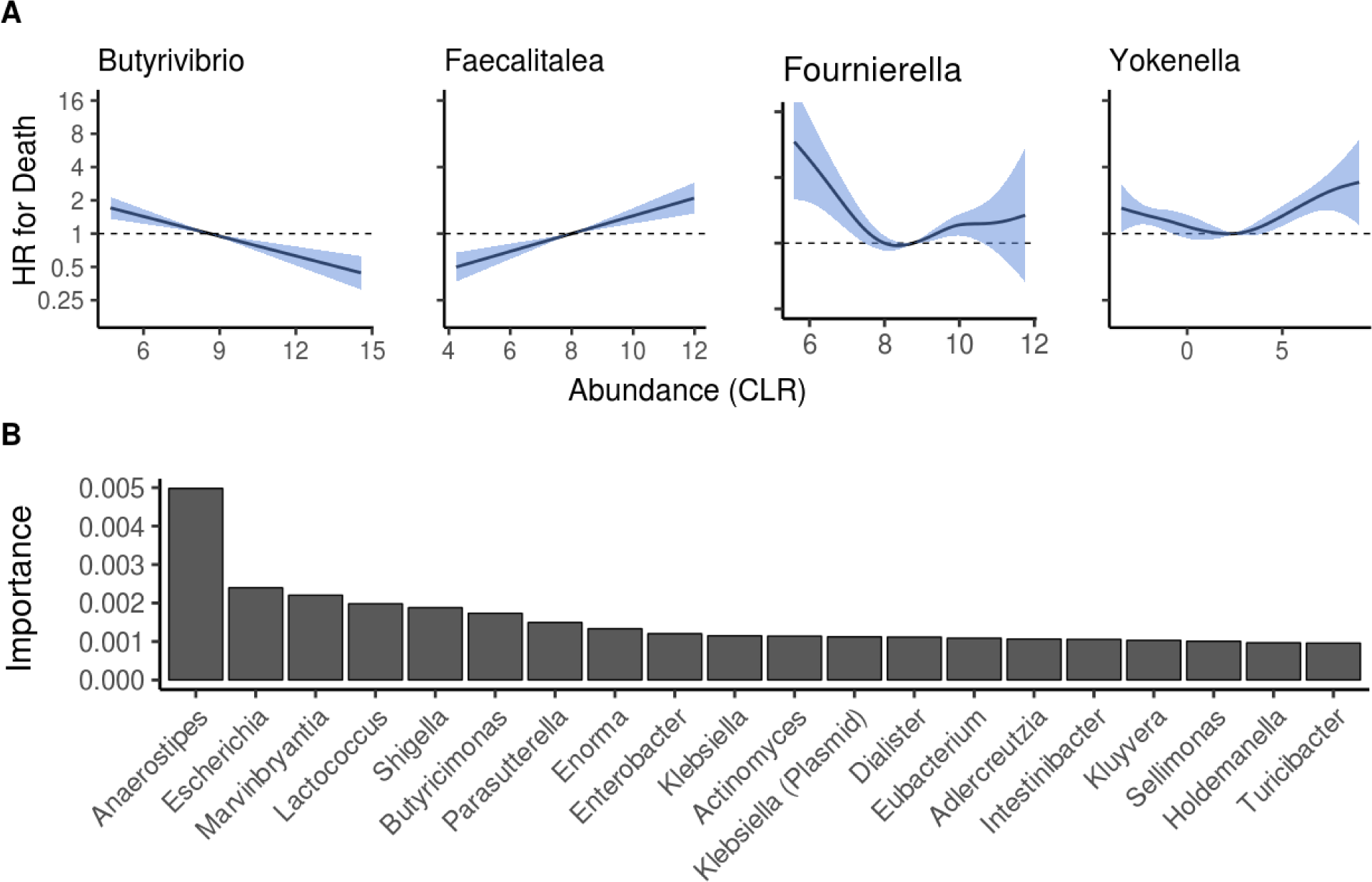
Relation of individual genera and core genera with mortality. **A** Examples of observed linear and non-linear associations between bacterial abundances and mortality. Blue area indicates the 95% confidence interval of the hazard ratio (HR). **B** Importance scores for top 20 genera based on multivariable random forest modeling. Analyses in panels A and B are adjusted for age, body mass index, sex, smoking, diabetes, use of antineoplastic and immunomodulating agents, systolic blood pressure and self-reported antihypertensive medication. Increased *Escherichia, Shigella* and *Kluyvera* were positively related to mortality, *Parasutterella* and *Faecalibacterium* were negatively associated with mortality, while this relation was nonlinear for *Phocea* and *Anaerostipes*.

**Extended Data Fig. 6.**
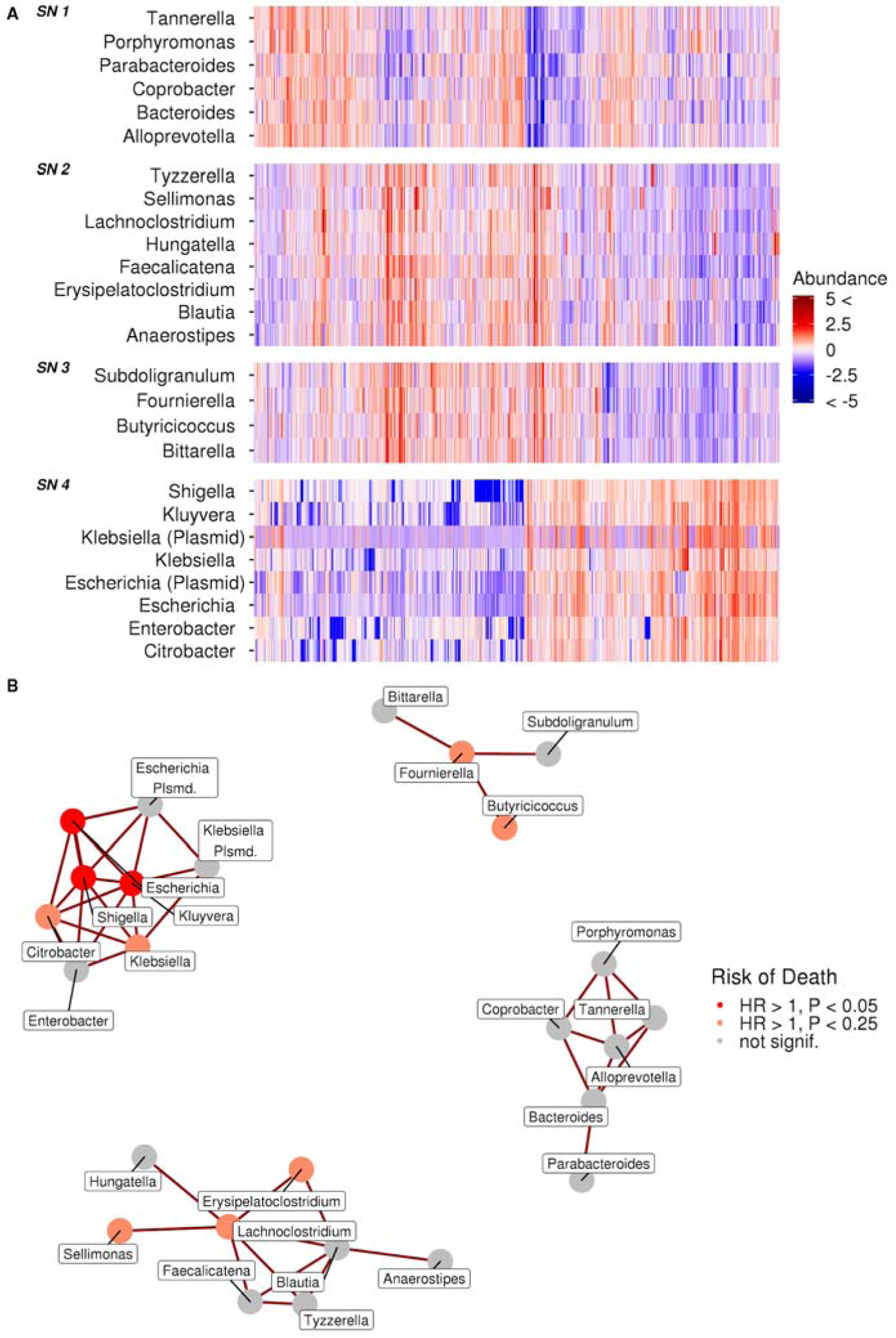
Microbial subnetworks and community structure. **A** Standardised log relative abundances of genera included in observed five microbial subnetworks across the study sample. We ordered the samples with hierarchical clustering (Ward method) based on Spearman correlation of CLR z-scores. **B** The observed network structure and the risk of death related to its components. HR, hazard ratio. Analyses are adjusted for age, body mass index, sex, smoking, diabetes, use of antineoplastic and immunomodulating agents, systolic blood pressure and self-reported antihypertensive medication; *P* values are FDR-adjusted. Subnetworks with only two or one genera not included in the plot.

**Extended Data Fig. 7.**
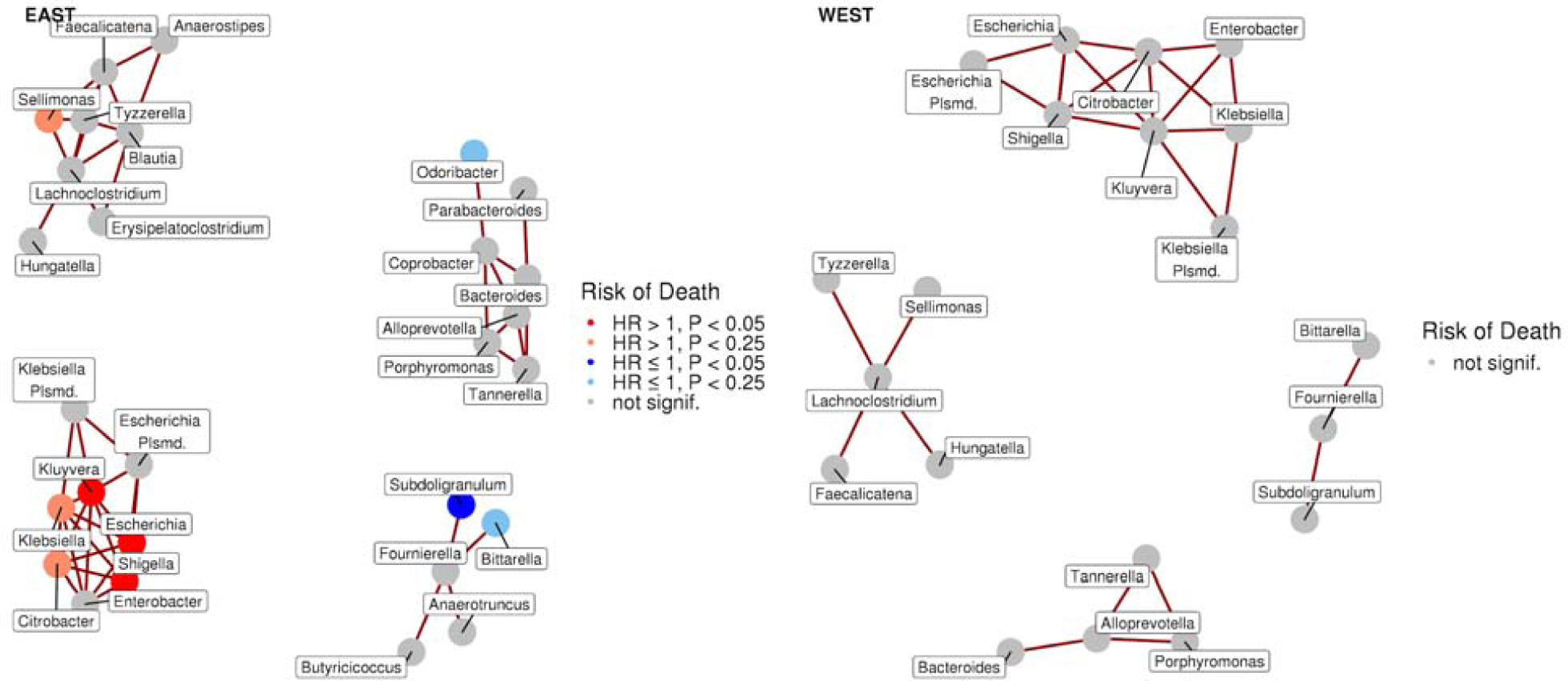
Network structure in Eastern and Western Finns. The mortality-associated taxonomic co-occurrence network (Escherichia, Shigella, Lambdavirus, Salmonella, and others) can be observed both in the Eastern (n=4871; 519 deaths) and Western (n=2184; 210 deaths) population with the identical procedure (SPIEC-EASI with the same parameter settings) as in the main analysis. Analyses are conducted separately for the Eastern and Western population and are adjusted for age, body mass index, sex, smoking, diabetes, use of antineoplastic and immunomodulating agents, systolic blood pressure and self-reported antihypertensive medication; *P* values are FDR-adjusted. Subnetworks with only two or one genera not included in the plot.

**Extended Data Fig. 8.**
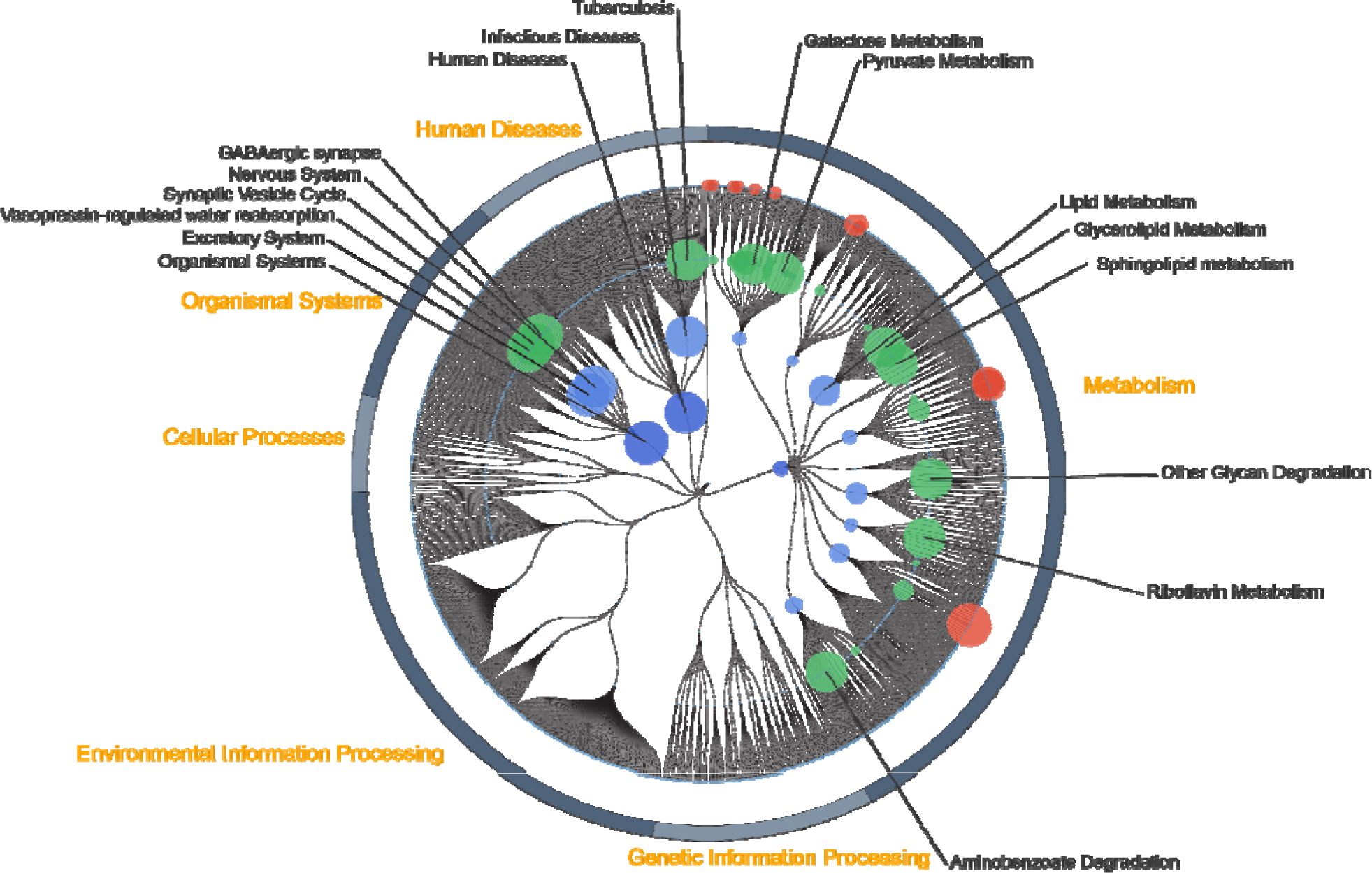
Functional pathways associated with decreased risk of death. For the module (red), pathway (green), biological process (light blue), and biological category (dark blue) functional layers, node size corresponds to the average inverse *P* value of the KEGG Orthology group assigned to that node. Only KO groups that were positively associated with mortality were included. Node titles are shown for nodes in the three highest layers with a size > 150.

## Supplementary Information

**Supplementary Table 1**. Distribution of phyla in FINRISK 2002.

<u>table_shogun_phylum_distribution.tsv</u>

**Supplementary Table 2**. Distribution of all and 1%-core genera in FINRISK 2002. The 1%-core microbiome is defined as the genera present with a within-sample relative abundance over 0.1% in at least 1% of samples.

<u>table_shogun_genus_distribution.tsv</u>

**Supplementary Table 3**. Significant relations of principal components 1–3 and alpha diversity with mortality.

<u>table_cox_diversities.tsv</u>

**Supplementary Table 4**. Top 20 drivers of the principal components 1–5.

<u>table_PC_drivers.tsv</u>

**Supplementary Table 5**. Association between individual genera and mortality

<u>table_cox_genus.tsv</u>

**Supplementary Table 6**. Prevalence of the VFDB virulence genes among the FINRISK samples.

<u>VFDB_Table.tsv</u>

**Supplementary Table 7**. Importance scores for core genera and covariates in random survival forest.

<u>table_random_survival_forest_importance.tsv</u>

**Supplementary Table 8**. Association between KEGG Orthology groups and mortality

<u>table_functional_cox.tsv</u>

**Supplementary Table 9**. SPIEC-EASI correlation coefficients between core genera.

<u>table_shogun_spieceasi_cor_coefficients.tsv</u>

